# Age-associated alterations in thalamocortical structural connectivity in psychosis-spectrum youths

**DOI:** 10.1101/2023.07.10.23292446

**Authors:** Lydia Lewis, Mary Corcoran, Kang Ik K. Cho, YooBin Kwak, Rebecca Hayes, Maria Jalbrzikowski

## Abstract

**Objective:** Psychotic symptoms typically emerge in adolescence when connections between the thalamus and cortex are still maturing. The extent to which thalamocortical connectivity differences observed in psychosis occur as a function of age-associated alterations is not fully understood.

**Methods:** We analyzed diffusion-weighted imaging data from 1254 participants 8-23 years old (typically developing youth: N=626, psychosis-spectrum youth: N=329, other psychopathology: N=299) from the Philadelphia Neurodevelopmental Cohort. Using deterministic fiber tractography, we modeled eight tracts between the thalamus and cortical regions of interest. We extracted diffusion spectrum imaging (DSI) and conventional diffusion tensor imaging (DTI) measures. We used generalized additive models to determine group and age-associated differences in thalamocortical connectivity.

**Results:** Compared to typically developing youth and youth with other psychopathologies, psychosis-spectrum youth exhibited thalamocortical reductions in DSI global fractional anisotropy (p-values range=3.0×10^−6^-0.05) and DTI fractional anisotropy (p-values range=4.2×10^−4^-0.03). Compared to typically developing youth, psychosis-spectrum youth exhibited shallower thalamus-prefrontal age-associated increases in DSI global fractional anisotropy and DTI fractional anisotropy during middle childhood, and steeper thalamus-prefrontal age-associated increases in those measures during adolescence. Both typically developing youth and youth with other psychopathologies exhibited decreases in mean and radial diffusivity in thalamus-frontal tracts during adolescence; psychosis-spectrum youth failed to show these age-related decreases.

**Conclusion:** Our findings suggest group differences and altered age-related patterns of thalamocortical white matter connectivity in psychosis-spectrum youth. Consistent alterations present as early as middle childhood and altered age-associated patterns during adolescence and young adulthood may contribute to the disruptions in thalamocortical connectivity observed in adults with psychosis.

## Introduction

Psychosis typically emerges in adolescence and early adulthood,^1^ a period characterized by ongoing refinement of connections between the cortex and subcortical structures, including the thalamus.^2–6^ In psychosis, thalamocortical connections are consistently disrupted across the phases of illness.^7–27^ However, the degree to which age-associated disruptions in thalamocortical connectivity contribute to psychosis remains unclear. Evaluating the age-associated patterns of thalamocortical structural connectivity in youth across the psychosis spectrum could help elucidate the neural basis of impairments associated with psychosis.

Studies of typical development find that white matter integrity in thalamocortical connections, as measured by fractional anisotropy (FA), increases with age.^3, 4, 28, 29^ In adults with schizophrenia, studies find reduced FA in white matter connectivity between the thalamus and prefrontal cortices^18, 19, 30–32^; some also find lower FA in the thalamus-occipital and thalamus-parietal connections.^30^ In a study that used probabilistic fiber tracking, in comparison to typically developing youth, psychosis-spectrum youth exhibited significantly lower FA values in tracts between the thalamus and six cortical regions (prefrontal, motor, somatosensory, temporal, posterior parietal and occipital cortices).^33^ This study found that typically developing and psychosis-spectrum youth exhibited linear age-related increases in FA between thalamus and motor and somatosensory cortices and linear age-related decreases in FA between thalamus and temporal and occipital cortices.^33^

These studies are foundational to our understanding of white matter alterations in psychosis, but we may learn more information by integrating recent developments in diffusion imaging methodology. First, most diffusion-weighted imaging studies of psychosis do not remove participants based on motion, though removing such poor quality scans influences age-associated patterns of neurodevelopment.^34^ Second, diffusion spectrum imaging (DSI) analyses improve upon the classical diffusion tensor imaging (DTI) model by allowing multiple directions of diffusion within a voxel.^35^ DSI methods lead to improved accuracy in mapping crossing fibers and enable the capture of information regarding tissue alterations that may affect the slow diffusion compartment^36–38^; thus, DSI is considered more sensitive to white matter microstructural organization than classical DTI. In DSI methodology, the Q-Space Diffeomorphic Reconstruction method^39, 40^ calculates quantitative anisotropy (QA), which measures anisotropic diffusion and incorporates spin density information.^41^ In one study, QA-based tractography outperformed FA-based tractography and produced the least number of false tracts when compared to other tractography-based approaches.^41^ Third, probabilistic tractography is the most common fiber tracking method used in psychosis studies.^42–46^ An alternative approach is deterministic tractography, which defines a white matter fiber trajectory beginning in a seed region and proceeding along the primary diffusion direction for each subsequent voxel. This method, unlike probabilistic tractography, does not include randomization but relies on local fiber orientation; thus, the computed trajectory from a given seed will always be the same.^47–49^ Furthermore, a recent study found that deterministic tractography detected 92% of valid connections while probabilistic tractography detected 45% of valid connections.^50^ Finally, many developmental changes that occur during adolescence are nonlinear^51–53^ and these patterns may be obscured when analyzed using a linear model. Flexible modeling approaches like generalized additive models (GAMs), which do not require a linear relationship between the predictor and the development variable, should be considered.

Leveraging these methodological advances, we used GAMs to 1) determine group differences in DSI and DTI measures extracted from thalamocortical connections in psychosis-spectrum youth vs. typically developing youth vs. youth with other psychopathologies, and to 2) examine the extent to which age-associated thalamocortical connectivity differed in psychosis-spectrum youth. We hypothesized that, similar to adults with schizophrenia,^18, 30–32^ psychosis-spectrum youth would show widespread group differences in DTI measures. Given the paucity of literature on DSI measures in psychosis, we did not have specific hypotheses about alterations in psychosis-spectrum youth. Finally, we hypothesized that when we used DSI measures of thalamocortical connectivity, we would observe age-associated alterations in psychosis-spectrum youth that may be obscured in DTI measures,^33^ and/or that the GAM models would capture distinct periods with specific age-associated alterations.

## Methods

### Pre-registration of investigation

We completed pre-registration of this project using the Open Science Framework website. [Details redacted for anonymization].

### Participants

The final neuroimaging dataset consisted of 1254 participants ages 8-23 years old from the Philadelphia Neurodevelopmental Cohort (PNC, Table 1). Inclusion criteria for PNC subjects included the ability to provide signed informed consent, English proficiency, and the ability to engage in psychiatric and cognitive phenotyping procedures. The subjects were recruited from a database at the Center for Applied Genomics at the [redacted for anonymization], as 78% of the 50,000 youths had provided consent to be re-contacted for future research. Exclusion criteria for the subset of subjects in the PNC who received neuroimaging included: medical problems that could impact brain function (such as severe medical problems, neurological conditions, or endocrine disorders), impaired vision/hearing, claustrophobia, or MRI contraindications.

**Table 1.** Participant baseline demographics.

|  | Typically<br>Developing | Other<br>Psychopathology | Psychosis-Spectrum |
| --- | --- | --- | --- |
| Total N | 626 | 299 | 329 |
| #F \| %F | 324 \| 51.76% | 176 \| 58.86% | 176 \| 53.5% |
| Mean Age | 15.38 | 14.91 | 15.89 |
| Age Range | 8.2-23 | 8.2-21.8 | 8.9-22.9 |

Abbreviations: F=female.

We used the GOASSESS, a modified version of the Kiddie-Schedule for Affective Disorders and Schizophrenia,^54^ to determine psychopathology history. Following completion of the GOASSESS, we defined psychosis-spectrum youth as participants who: 1) had a score of 6 on any PRIME Screen Revised item; had a score of 5 or 6 on three or more items on the PRIME Screen Revised; or scored 2 standard deviations or more above the total score of age-cohort mean on the SIPS; or 2) answered ‘yes’ to hallucination related questions on the KSADS, reported that they were not using drugs at the time the symptom was experienced, and endorsed experiencing significant impairment or distress as a result; or 3) scored 2 standard deviations or more above the age-cohort mean total score on six SOPS negative symptom items: attention and focus, disorganized speech, perception of self, experience of emotion, occupational function and avolition. We defined typically developing youth as youth who denied clinically significant symptoms of psychopathology based on responses to the GOASSESS interview. To define the “other psychopathology” group, we used responses to questions on the GOASSESS to determine DSM-IV diagnosis ranking. Like other PNC publications,^55, 56^ we considered psychopathology to be significant if symptoms endorsed were consistent with frequency and duration of a DSM-IV psychiatric disorder, while correspondingly accompanied by significant distress or impairment (rating of >5 on a scale of 0-10).

### MRI Acquisition

All PNC scans were collected with a 32-channel head coil on a single 3T Siemens Tim Trio whole-body scanner at the Hospital of the University of Pennsylvania. Diffusion-weighted imaging was split into two separate imaging runs, with a full scanning time of approximately 11 minutes. Scans were acquired with the following parameters: TR=8100 ms, TE=82 ms, FOV=240 by 240 mm; Matrix=128 x128 x70, in-plane resolution=1.875mm2; slice thickness=2 mm, gap=0 mm; FlipAngle=90°/180°/180°, volumes=71 (35 in first run, 36 in second run), GRAPPA factor=3, bandwidth=2170 Hz/pixel, PE direction=AP. The DWI sequence was a twice-refocused spin-echo (TRSE) single-shot EPI sequence, consisted of 64 b=1000s/mm2 diffusion-weighted volumes and 7 b=0s/mm2 volumes. In order to minimize eddy current artifacts,^57^ a four-lobed diffusion encoding gradient scheme and a 90-180-180 spin echo sequence were used. For more details see Satterthwaite et al., 2014.^58^

### MRI Processing

We merged two separate runs for each subject into a single data set (71 volumes). We performed a Gibbs ringing artifact removal^59^ and then Eddy current-induced distortion and motion correction using Eddy, from FSL, with outlier removal function.^60, 61^ We then used the q-space diffeomorphic reconstruction (QSDR) method, available in the DSI Studio Software,^62^ to reconstruct the images and warp them to standard MNI space.

### Thalamic Region of Interest

We created the thalamic region of interest (ROI) using the Morel Thalamic Atlas that was adapted to 3D MNI space.^63^ We merged the individual thalamic nuclei to create the whole thalamus (i.e., merged anterior, mediodorsal, lateral geniculate, ventral anterior, ventral lateral, ventral posterior lateral, ventral posterior medial, pulvinar, lateral dorsal, lateral posterior, and intralaminar regions). See Figure 1A for a visual depiction.

**Figure 1.**
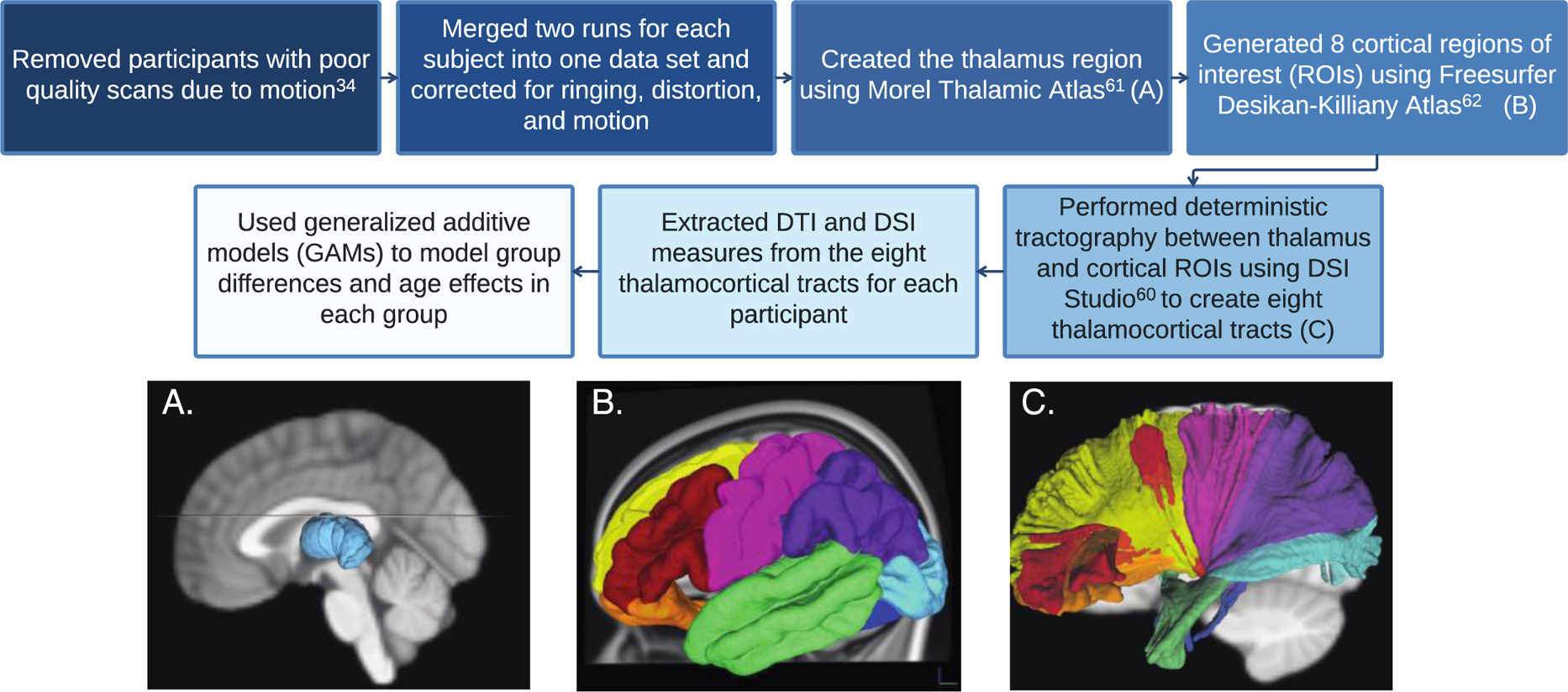
Diffusion weighted imaging analysis pipeline. **A.** Thalamic region of interest (light blue). **B.** Cortical regions of interest: the lateral prefrontal cortex (red), medial prefrontal cortex (yellow), orbitofrontal cortex (orange), somatomotor cortex (pink), lateral temporal cortex (green), medial temporal cortex (dark blue), parietal cortex (purple), occipital cortex (light blue). **C.** Tracts between the thalamus and the lateral prefrontal cortex (red), medial prefrontal cortex (yellow), orbitofrontal cortex (orange), somatomotor cortex (pink), lateral temporal cortex (green), medial temporal cortex (dark blue), parietal cortex (purple), occipital cortex (light blue).

### Cortical Parcellation

Similar to previous publications on thalamocortical connectivity,^14, 19^ we generated eight cortical ROIs (lateral prefrontal cortex, medial prefrontal cortex, orbitofrontal cortex, somatomotor cortex, lateral temporal cortex, medial temporal cortex, parietal cortex, and occipital cortex) using the FreeSurfer Desikan-Killiany atlas^64^ as reported in Supplementary Table 1 and shown in Figure 1B. Because we did not have any a priori hypotheses about laterality, we calculated bilateral connections between the thalamus and all cortical regions.

### Tractography

We performed deterministic tractography using DSI Studio.^62^ We performed the first set of tracking between the bilateral whole thalamus and each bilateral cortical ROI using the Human Connectome Project group average template image of 1021 subjects.^65^ We designated both the thalamus and the selected cortical region as ROIs and each of the seven other cortical regions as regions of avoidance. After performing tractography, we manually edited tracts to remove any fibers that strongly deviated.

We performed tracking with a threshold QA of 0.1, an angular threshold of 60 degrees, and a 0.5 mm step size (half the size of one voxel). We used 30 mm as the minimum length and 120 mm as the maximum length and removed any tracts falling outside of these values. We terminated tracking after 5,000,000 seeds. We selected tractography parameters similar to previous publications that used deterministic QA-based tractography in DSI Studio.^66–69^ After tractography, we converted these eight thalamocortical tracts into ROIs and extracted DSI values from each subject for each of the eight tracts.

### DTI/DSI Measures

Extracted DTI and DSI measures from the eight thalamocortical tracts are described below.

### DTI measures

DTI measures assess the diffusion signal by using a tensor model.^70, 71^ Fractional anisotropy (FA) estimates the anisotropy of diffusion of water molecules across a tissue and ranges between 0 (completely isotropic) and 1 (completely anisotropic) and is considered an overall measure of white matter integrity.^72, 73^ Axial diffusivity (AD) indicates the rate at which molecules diffuse in the primary diffusion direction and is associated with axonal density.^74–76^ Radial diffusivity (RD) measures the rate at which water molecules diffuse perpendicular to the primary diffusion direction and is associated with myelination.^74, 77, 78^ Mean diffusivity (MD) measures the average amount of diffusion in a voxel and reflects the amount of water in the extracellular space.^79, 80^

### DSI Measures

Diffusion spectrum imaging analyses (DSI) improve upon the classical diffusion tensor model by allowing for the possibility of multiple directions of diffusion within a single voxel.^35^ DSI measures are derived from a model-free approach that uses information from a diffusion orientation distribution function; evidence suggests these measures better represent complex fiber organization.^41, 81^ Global fractional anisotropy (GFA) is considered the DSI analogue to FA and is believed to reflect white matter integrity. GFA ranges between 0 and 1 and corresponds to the magnitude of the principal diffusion direction in each voxel.^81^ Quantitative anisotropy (QA) is a measure of anisotropic diffusion that incorporates spin density information^41^ and is associated with axonal density.^82, 83^ Isotropy (ISO) measures background isotropic diffusion and is believed to reflect cerebrospinal fluid and edema.^82^ Restricted diffusion imaging (RDI) indicates the total amount of restricted diffusion in a tissue in any orientation and is believed to reflect cell density and inflammation.^84^

### Image Quality Assessment

To assess image quality and remove participants with poor quality scans due to motion, we calculated a temporal signal-to-noise ratio (TSNR), based on a previous publication.^34^ We estimated TSNR at each brain voxel for the 64 b=1000 s/mm2 DTI volumes. We then obtained a single SNR measure by averaging all brain voxel TSNRs. We chose this measure to assess image quality because it differentiated poor data from usable data with a high degree of accuracy.^34^ We used the defined cut-off point of 6.47 to exclude poor quality scans from our analyses.^34^

### Statistics

We used general additive models (GAMs) to examine group differences (typically developing vs. psychosis spectrum vs. other psychopathology) and smoothed age effects in each group on all DTI and DSI measures. A GAM is an extension of the general linear model but does not assume a linear relationship between the predictor and dependent variable, allowing for a more flexible relationship. To avoid overfitting, GAMs assess a penalty on nonlinearity. Smoothed predictor functions are automatically derived during model estimation with basis functions. Age is modeled as a smooth function (continuous derivatives), which is akin to the “age-related” slope in a general linear model. Because the relationship between the smoothed predictor and the dependent variable is not required to have the same functional form in each group, we were able to examine the smoothed effects of chronological age for all groups separately. We included sex in all models as a covariate. Like other non-parametric approaches, GAMs may be sensitive to outliers; we attempted to protect against this limitation by limiting the number of splines that can occur when the line of best fit is determined (a maximum of three knots), and we used restricted maximum likelihood to optimize the smoothness fit. After running all models, we used False Discovery Rate to correct for multiple comparisons and obtain a corrected p-value (i.e., q-value).

To determine time periods in which significant change was occurring in each group, we used a multivariate normal distribution whose vector of means and covariance were defined by the fitted GAM parameters to simulate 10,000 GAM fits and their first derivatives, generated at 0.1-year age intervals. Similar to previous publications^85–87^ and in line with recent guidelines,^88^ we defined significant intervals of age-related change in MRI measures as ages when the 95% confidence intervals of simulated GAM fits did not include zero.

To determine time periods during which age effects in each group differed from another group (e.g., typically developing age effects vs. psychosis spectrum age effects), we took the difference between the upper and lower 95% confidence intervals of the smoothed fit in two groups, henceforth called the ‘difference in smooths’. For each dependent variable, we considered effects of age to be significantly different in the two groups being compared during periods of time in which the difference in smooths did not include zero. This approach has been used in previous publications.^87, 89, 90^

## Results

Group differences, age effects, and differences in age-related slopes between groups are reported in Table 2. In the main text, we report on statistically significant group differences and periods of time when age-related smooths differed between typically developing youth and psychosis-spectrum youth (i.e., ‘differences in smooths’). We also report when group and age-related differences were present in psychosis-spectrum youth vs. youth with other psychopathologies. Discussion of descriptive statistics (i.e., time periods of age-related changes within a specific group) and periods of time when age-related slopes differed only between psychosis-spectrum youth and youth with other psychopathologies are reported in the Supplemental Text.

**Table 2.**
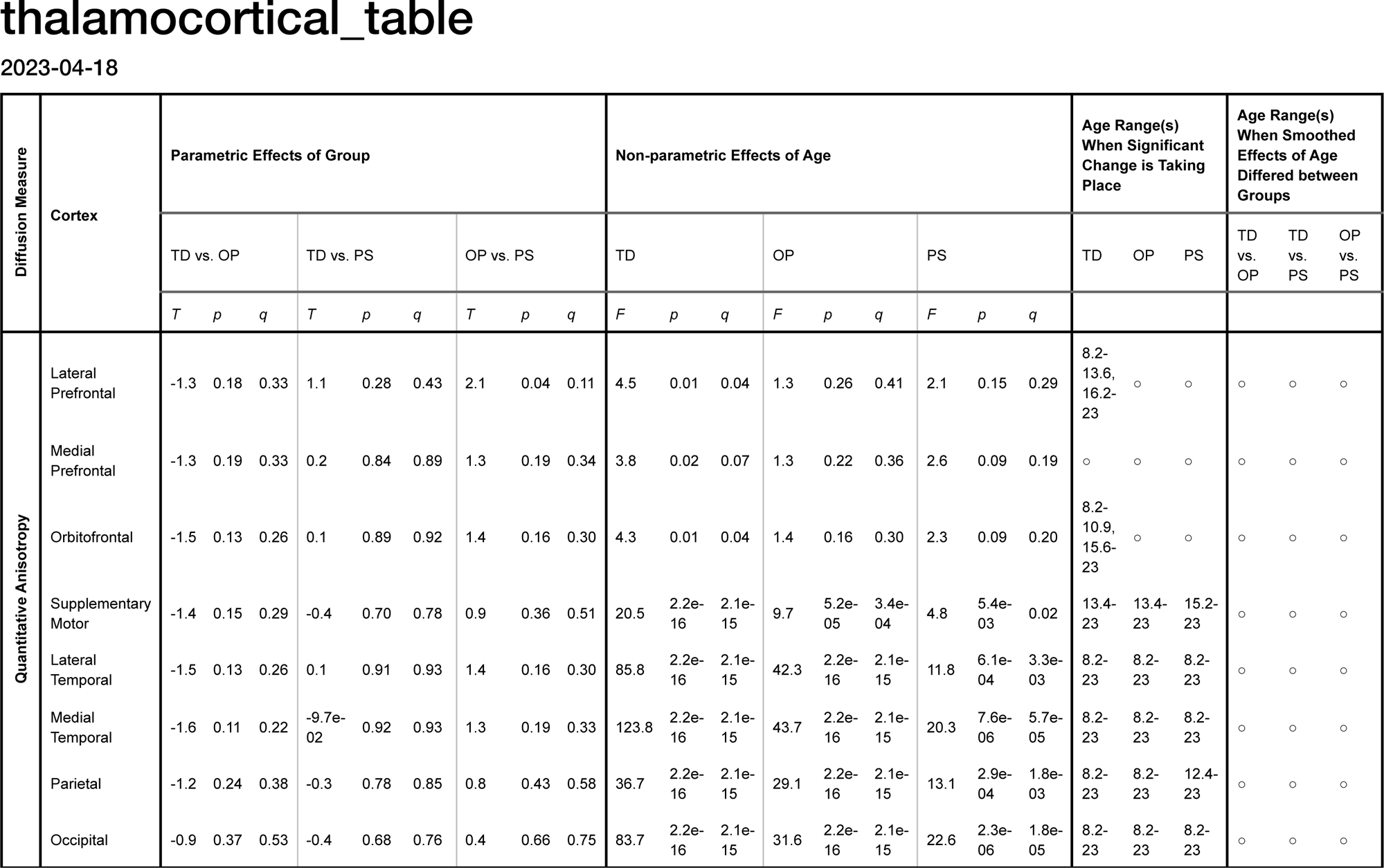
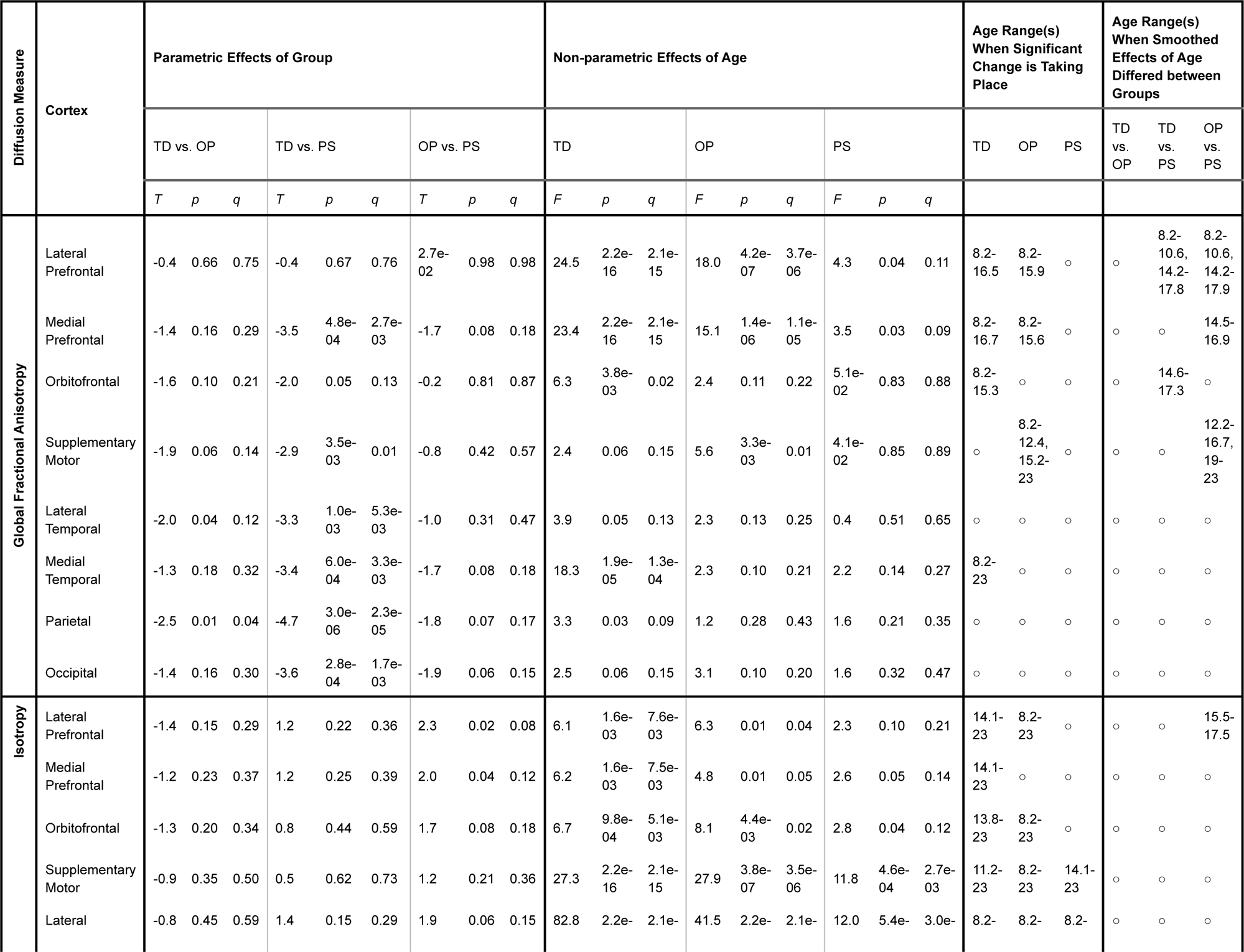
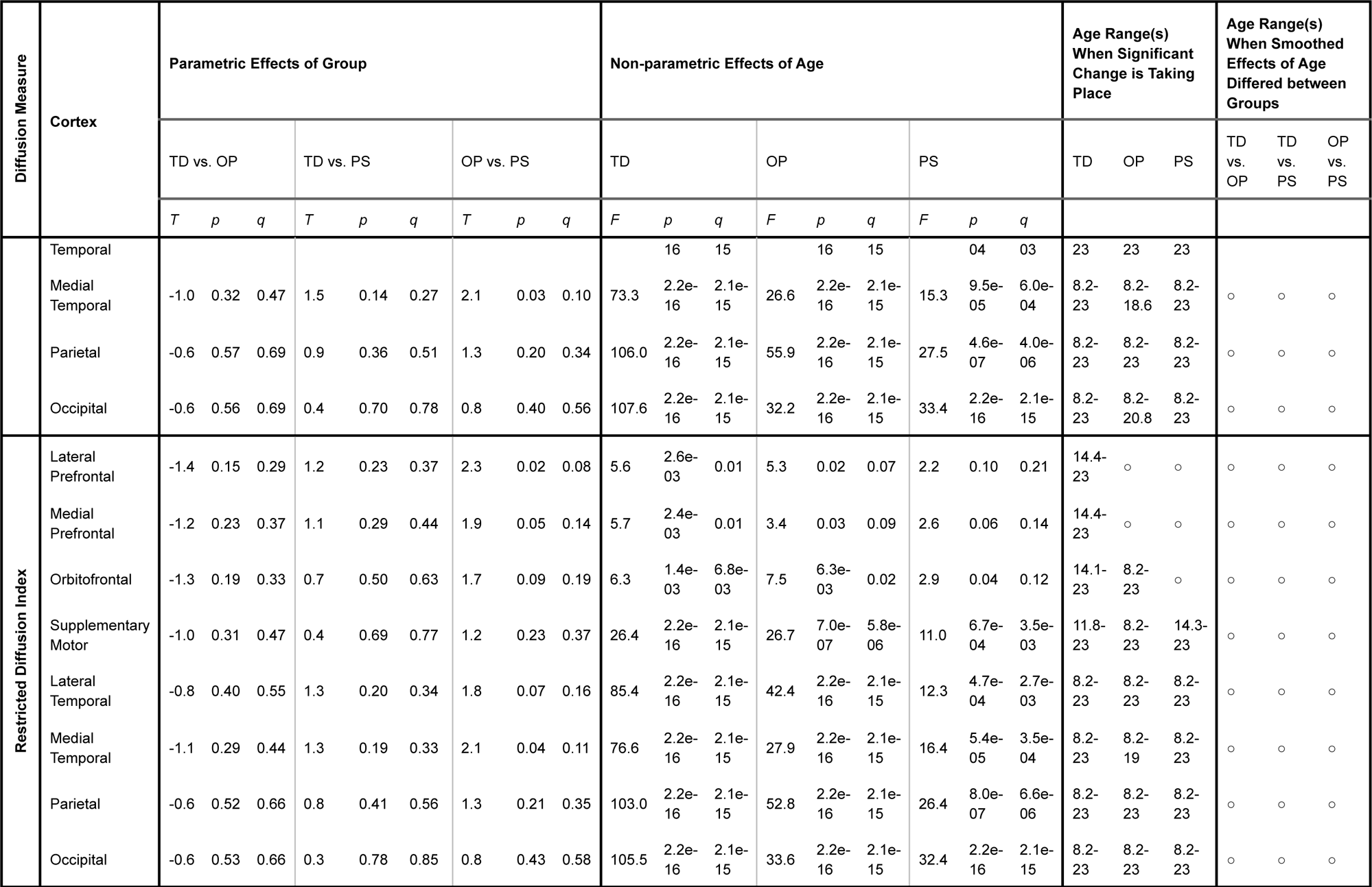
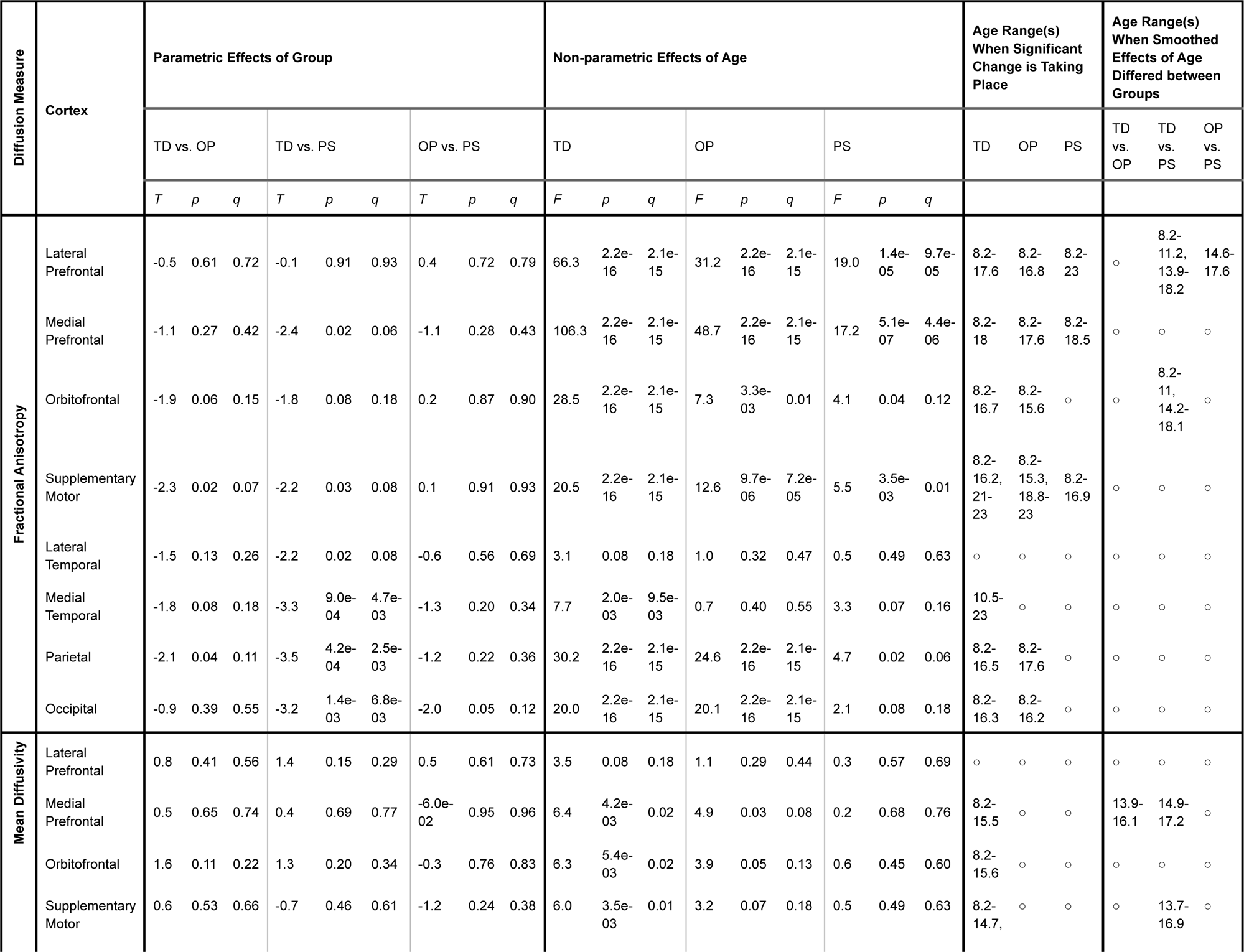
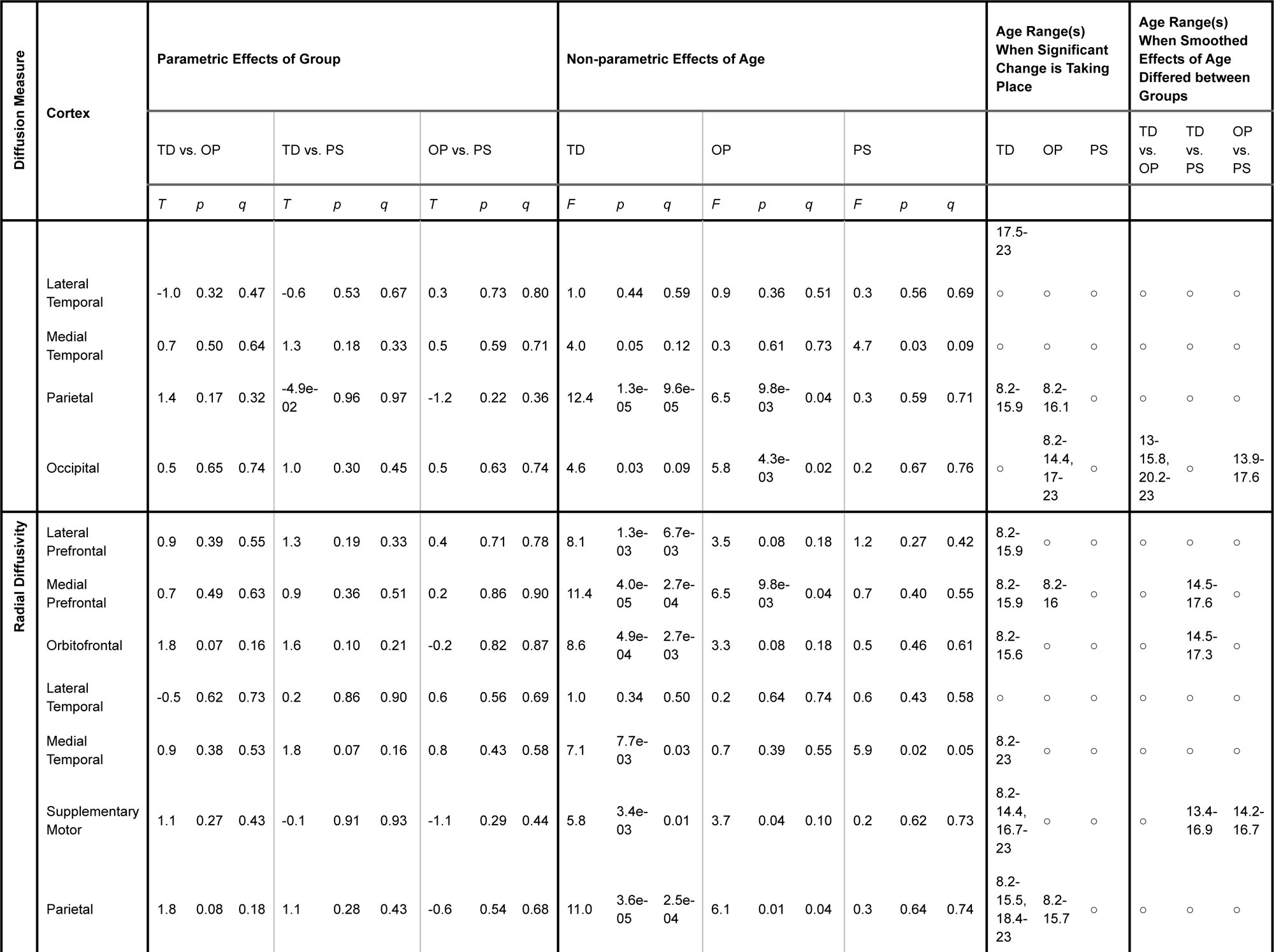
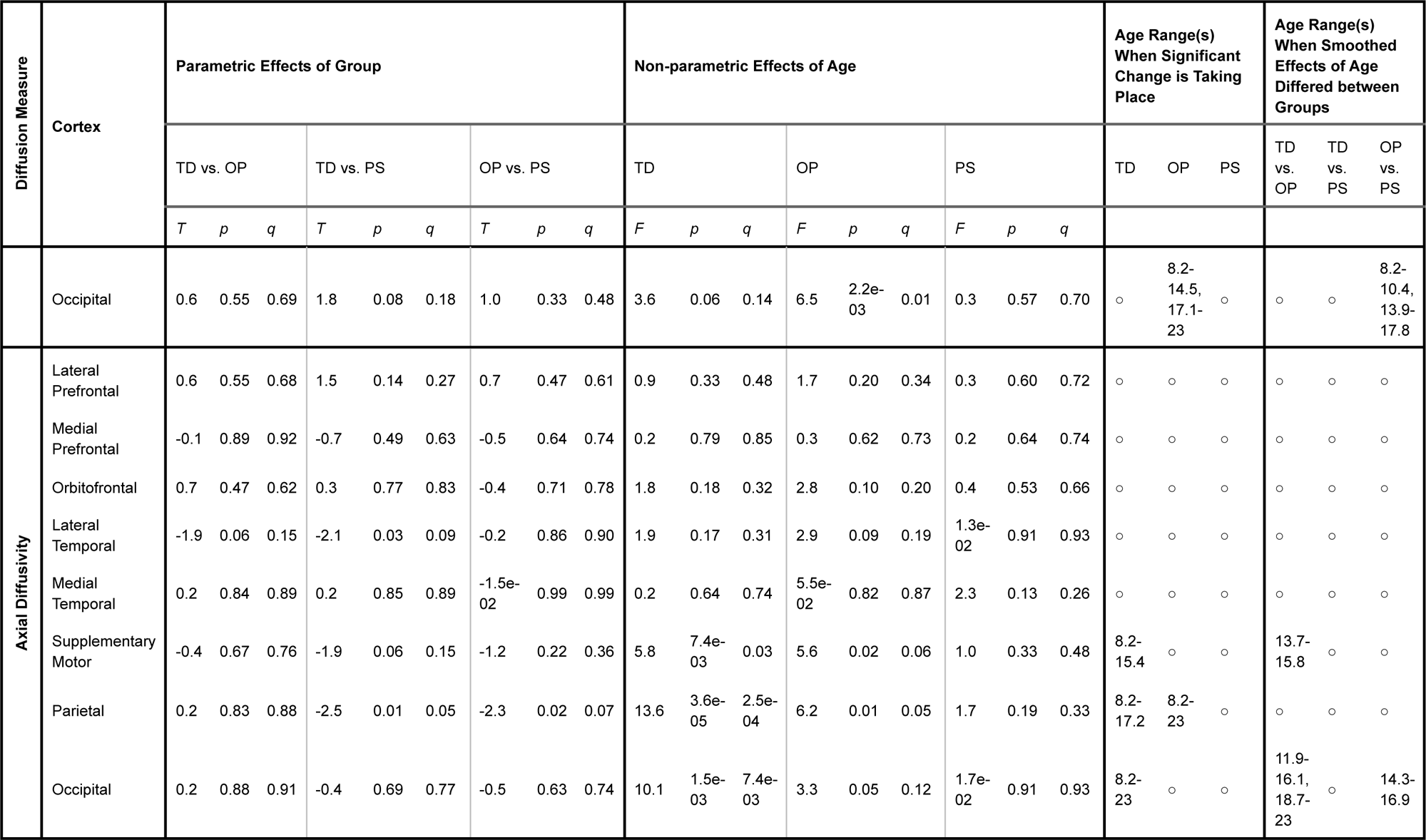
Group and Age Effects on Thalamocortical Trajectories in Psychosis-spectrum youth vs. Typically Developing Youth vs. Youth with Other Psychopathologies: DSI and DTI Measures.

### Psychosis-spectrum youth have reductions in GFA and FA thalamocortical connectivity

In comparison to typically developing youth, psychosis-spectrum youth exhibited reduced GFA in connections between the thalamus and the following cortices: medial prefrontal, somatomotor, lateral temporal, medial temporal, parietal, and occipital (all *q*<.01, Table 2, Figure 2A-H). Youth with other psychopathologies had lower thalamus-parietal cortex GFA in comparison to typically developing youth (*T*=-2.5,*p*=0.01,*q*=0.04), but not psychosis-spectrum youth (T=1.8,p=0.07,q=0.17). Youth with other psychopathologies did not exhibit disruptions in GFA when compared to either group (all *q*>.05) in any other thalamocortical tract.

**Figure 2.**
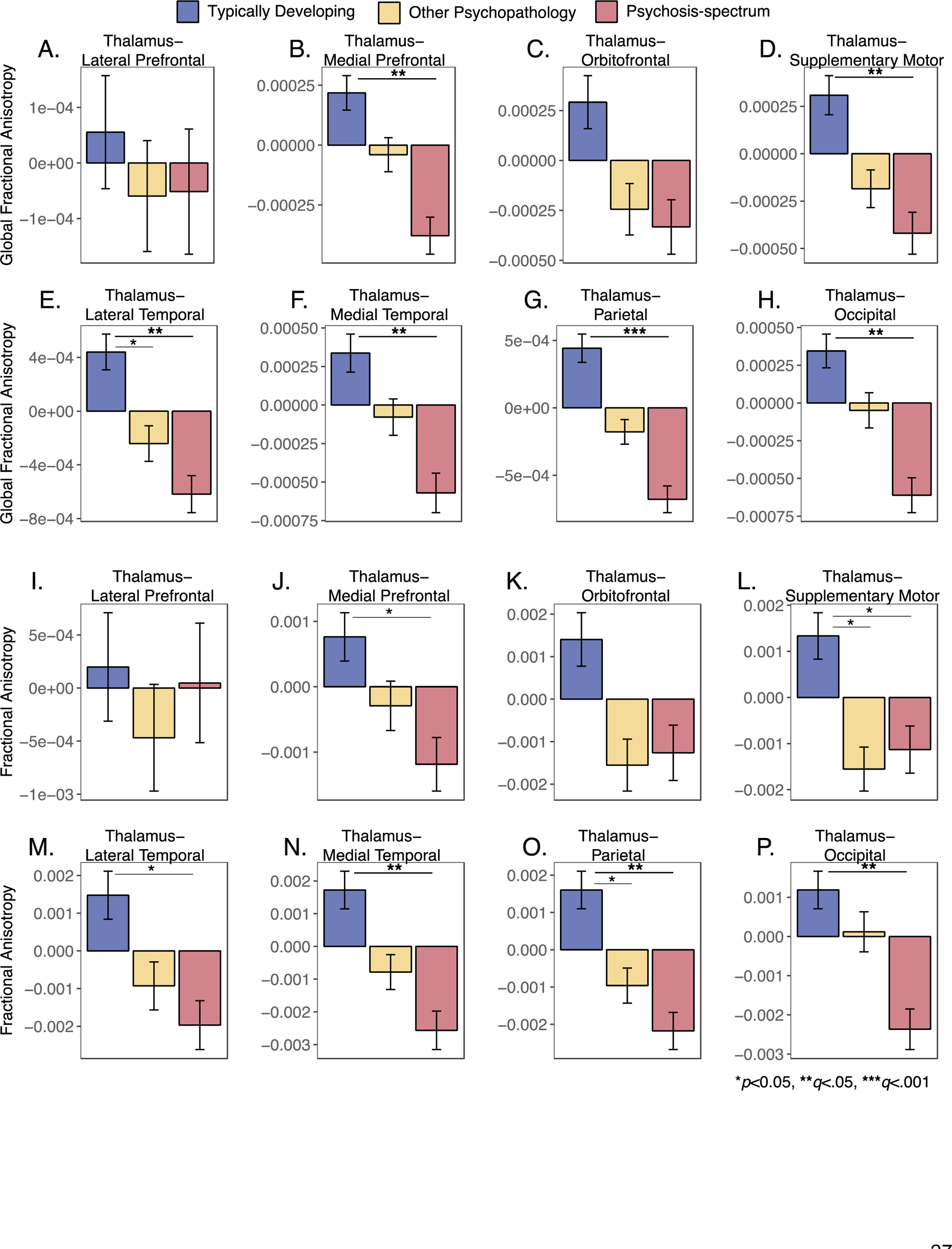
**A.-H.** Group differences in thalamocortical global fractional anisotropy in Typically Developing Youth (blue) vs. Youth with Other Psychopathologies (yellow) vs. Psychosis-spectrum youth (red). **I.-P.** Group Differences in thalamocortical fractional anisotropy in Typically Developing Youth (blue) vs. Youth with other Psychopathologies (yellow) vs. Psychosis-spectrum youth (red).

Compared to typically developing youth, psychosis-spectrum youth exhibited reductions in thalamus-medial temporal, thalamus-parietal, and thalamus-occipital FA (all q<.0068, Table 2, Figure 2I-P). Youth with other psychopathologies did not differ from typically developing or psychosis-spectrum youth on any FA measures (all *q*>.07). Psychosis-spectrum youth exhibited reduced thalamus-parietal AD in comparison to typically developing youth (*q*=.05, Table 2). Youth with other psychopathologies youth did not exhibit significant differences in AD when compared to either group (all *q*>.07).

### Psychosis-spectrum youth have age-associated alterations in prefrontal thalamocortical FA and GFA

Both typically developing youth and youth with other psychopathologies exhibited age-associated increases in thalamus-lateral prefrontal GFA from 8.2-16 years. Difference in smooths revealed that, in comparison to typically developing youth and youth with other psychopathologies, from 8-10 years old, psychosis-spectrum youth showed a shallower age-related increase in thalamus-lateral prefrontal GFA. However, during the teenage years (14.2-17.8 years), psychosis-spectrum youth exhibited a steeper age-related increase in thalamus-lateral prefrontal GFA in comparison to typically developing youth and youth with other psychopathologies (Figure 3A). Typically developing youth also exhibited an age-associated increase in thalamus-orbitofrontal GFA from 8.2-15.3 years. Psychosis-spectrum youth and youth with other psychopathologies did not exhibit an age associated effect in this tract. Difference in smooths revealed that psychosis-spectrum youth failed to exhibit the inverted u-shaped age-related slope that typically developing youth exhibited during adolescence (14.6-17.3 years) in thalamus-orbitofrontal GFA (Figure 3C). Derivate plots in Figure 3C show that, during this time, typically developing youth exhibited a significant increase and a subsequent decrease in thalamus-orbitofrontal GFA, but psychosis-spectrum youth do not show any age-associated effects.

**Figure 3.**
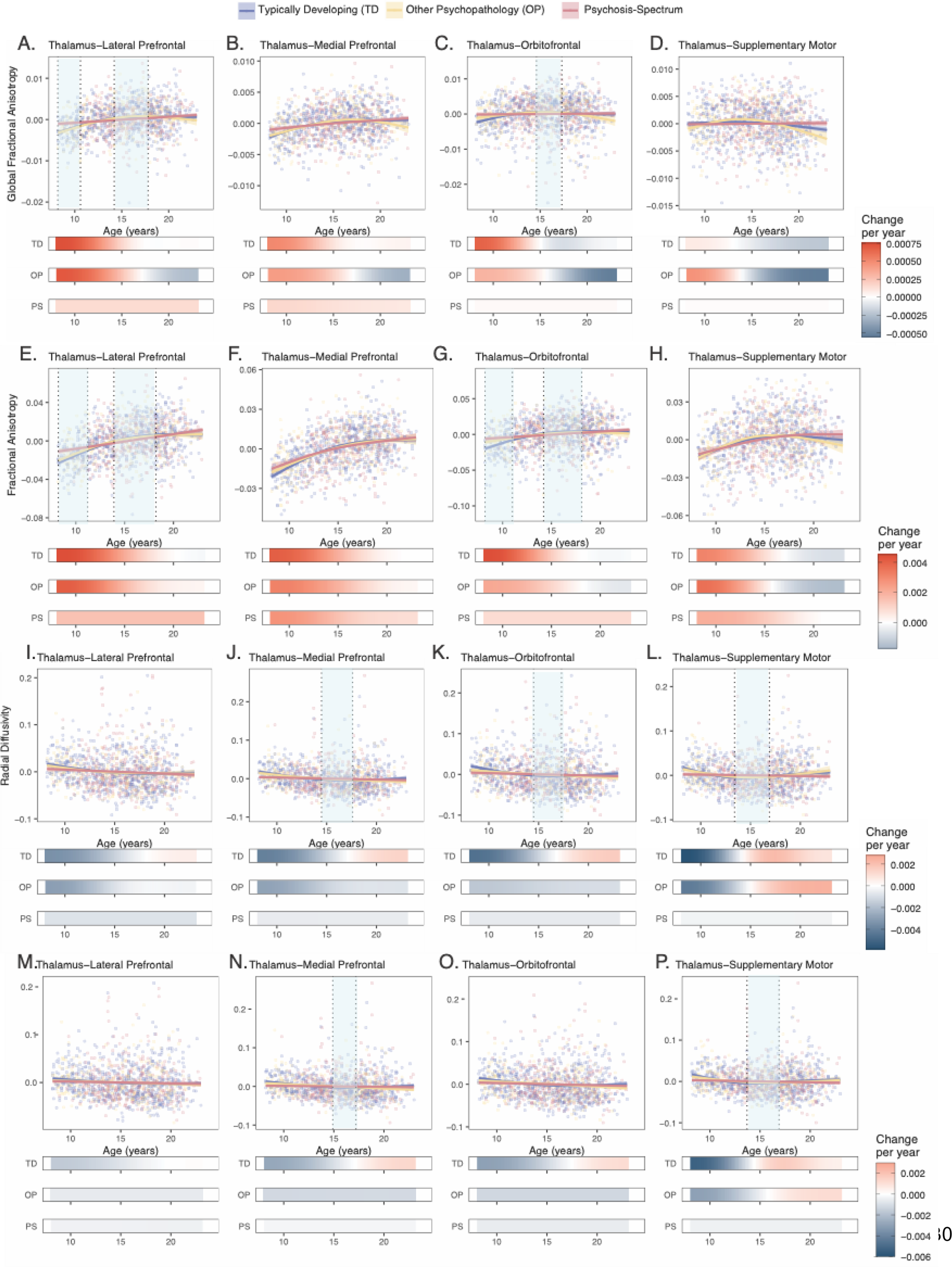
Neurodevelopmental trajectories of global fractional anisotropy, fractional anisotropy, radial diffusivity, and mean diffusivity for psychosis-spectrum youth, typically developing youth, and youth with other psychopathologies. Partial residual plots of A-D global fractional anisotropy trajectories, E-H fractional anisotropy trajectories, I-L radial diffusivity trajectories, and M-P mean diffusivity trajectories in tracts between the thalamus and the lateral prefrontal cortex (A, E, I, M), medial prefrontal cortex (B, F, J, N), orbitofrontal cortex (C, G, K, O), somatomotor cortex (D, H, L, P) for typically developing youth (blue) youth with other psychopathologies (yellow) and psychosis-spectrum youth (red). The partial residual plots reflect the relationship between age and the respective neuroimaging measures, given the covariates in the model. For each group, the thick line reflects the line of best fit. The bars underneath the age plots reflect the derivative of the slope, i.e., the rate of change taking place at a particular age. Darker blue indicates that there is a stronger decrease in the respective DSI or DTI measure taking place at that age, while brighter red indicates a stronger increase in the respective DSI or DTI measure. Dotted lines and areas of lighted shaded blue indicate times when there was a significant “difference in smooths” between Typically Developing and Psychosis Spectrum youth.

We observed a similar pattern of results for thalamus-prefrontal FA measures. Typically developing youth and youth with other psychopathologies exhibited age-associated increases in thalamus-lateral prefrontal FA from 8.2-17 years, and psychosis-spectrum youth showed an age-associated increase in thalamus-lateral prefrontal FA from 8.2-23 years. Difference in smooths revealed that, in comparison to typically developing youth, from 8-11 years old, psychosis-spectrum youth showed a shallower age-related increase in thalamus-lateral prefrontal FA. However, during the teenage years (14-18 years), psychosis-spectrum youth exhibited a steeper age-related increase in thalamus-lateral prefrontal FA in comparison to both typically developing youth and youth with other psychopathologies (Figure 3E). Typically developing youth and youth with other psychopathologies also exhibited an age-associated increase in thalamus-orbitofrontal FA from 8.2-16 yearsPsychosis-spectrum youth did not exhibit an age associated effect in this tract. Difference in smooths revealed that, in comparison to typically developing youth, from 8-11 years old, psychosis-spectrum youth showed a shallower age-related increase in thalamus-orbitofrontal FA. However, during the teenage years (14-18 years), psychosis-spectrum youth exhibited a steeper age-related increase in thalamus-orbitofrontal FA in comparison to typically developing youth (Figure 3G). Psychosis-spectrum youth did not show an altered age-related increase in thalamus-orbitofrontal FA in comparison to youth with other psychopathologies.

### Psychosis-spectrum youth exhibit age-related alterations in thalamus-frontal RD

Typically developing youth exhibited age-associated decreases in thalamus-medial prefrontal and thalamus-orbitofrontal RD from middle childhood through adolescence. Psychosis-spectrum youth did not show a significant effect of age. Difference in smooths revealed that, in comparison to typically developing youth, psychosis-spectrum youth failed to show the u-shaped age-related effect (i.e., a significant decrease in RD followed by a significant increase in RD) during adolescence (14-17 years) in thalamus-medial prefrontal and thalamus-orbitofrontal RD (Figure 3J-K). For RD in the thalamus-somatomotor tract, typically developing youth exhibited a u-shaped age effect, with significant *decreases* in RD from 8-14 years old, followed by age-associated *increases* from 16-23 years (Figure 3L). Psychosis-spectrum youth and youth with other psychopathologies failed to exhibit any significant age effects. Differences in smooths revealed that from ∼13-17 years old, psychosis-spectrum youth failed to show the u-shaped age-associated effect exhibited by typically developing youth and youth with other psychopathologies (first a steeper decrease, followed by a steeper increase) in thalamus-somatomotor RD (Figure 3L).

### Psychosis-spectrum youth do not exhibit typical age-related decreases in prefrontal MD

Typically developing youth exhibited significant decreases in thalamus-medial prefrontal MD tracts from 8-15 years, while psychosis-spectrum youth and youth with other psychopathologies did not show this effect. Difference in smooths revealed that psychosis-spectrum youth failed to show the u-shaped age-associated effect that typically developing youth exhibited during adolescence (14.9-17.2 years) in thalamus-medial prefrontal MD (Figure 3N). Derivative plots in Figure 3N show that with increasing age during this period, typically developing youth exhibited significant decreases in MD, followed by significant increases, while psychosis-spectrum youth did not. A similar pattern was observed between typically developing youth and psychosis-spectrum youth in thalamus-somatomotor MD between 13.7-16.9 years of age (Figure 3P).

Psychosis-spectrum youth and typically developing youth did not exhibit age-related differences in slopes in measures of QA, ISO, RDI, or AD.

## Discussion

We assessed age-related patterns in structural connectivity between the thalamus and the cortex in typically developing youth, psychosis-spectrum youth, and youth with other psychopathologies. In comparison to typically developing youth and youth with other psychopathologies, psychosis-spectrum youth exhibited widespread thalamocortical reductions in measures of DSI GFA and DTI FA. Compared to typically developing youth and youth with other psychopathologies, psychosis-spectrum youth exhibited a significantly shallower slope in thalamus-prefrontal GFA and FA and in middle childhood (∼8-10 years) but a steeper age-associated slope in adolescence (14-17 years). Psychosis-spectrum youth also failed to exhibit age-associated decreases in thalamus-prefrontal mean diffusivity and thalamus-somatomotor radial diffusivity, patterns that were observed in typically developing youth and youth with other psychopathologies. Our results provide a novel view of developmental alterations in thalamocortical white matter connectivity in youth experiencing psychosis spectrum symptoms, implicating differences in age-associated slopes at discrete periods of development in thalamocortical connections that are implicated in a wide array of sensory and cognitive processes.

Previous studies of adults with psychosis have found widespread lower FA across white matter tracts.^14, 15, 18–20, 91^ We found similar results in psychosis-spectrum youth with DTI FA measures and with a DSI analogue, GFA. In a study that used the same sample but used DTI FA and probabilistic tractography, researchers found strikingly similar results with regards to DTI FA: psychosis-spectrum youth exhibited lower FA levels across all thalamocortical tracts in comparison to typically developing youth.^33^ Taken together, these findings suggest widespread reductions in white matter integrity that are not driven by the issues associated with the tensor model, such as crossing fibers; furthermore, these reductions are robust to methodological differences. This consistent reduction in GFA and FA in multiple brain regions may indicate global deficits rather than local deficits. Previous studies used alternative analytical approaches based on systematic resampling of an original data set, supporting that global, and not just local, fractional anisotropy deficits are characteristic of psychosis.^92^ Taken together, these findings suggest that features of psychosis are caused by subtle, widespread disruptions in white matter connectivity. Additionally, our findings indicate that thalamocortical white matter disruptions may be present in early stages of psychotic illness and are observed in individuals as young as 8 years old with sub-threshold and full-blown psychotic symptoms.

Our findings suggest that age-associated changes in white matter connectivity, as measured through GFA and FA, may be disrupted throughout adolescence in psychosis-spectrum youth. Specifically, during middle childhood, psychosis-spectrum youth exhibited a significantly shallower slope in thalamus-prefrontal GFA and FA in comparison to typically developing youth and youth with other psychopathologies. This may mean that psychosis-spectrum youth who have onset of symptoms during middle childhood have neural circuitry that is more strongly affected. Alternatively, it is possible that psychosis-spectrum youth had “precocious” decreases in connectivity that occurred prior to the age range we studied. In support of this hypothesis, there is some evidence that the brains of individuals with psychosis “age” faster.^93^ During adolescence, psychosis-spectrum youth exhibited a steeper age-associated slope in thalamus-lateral prefrontal and thalamus-orbitofrontal tracts than typically developing youth and youth with other psychopathologies.

Adolescence has been conceptualized as a period of specialization when cellular mechanisms and neural circuitry motivate experience-seeking behaviors^94^ which in turn stimulate experience-dependent plasticity, strengthening neural synchrony and refining cortical networks.^94^ If the thalamus-prefrontal GFA and FA trajectories of typically developing youth during adolescence represent a period of specialization (when higher-level systems that contribute to adult outcomes are formed^86, 95^), an abnormal trajectory during middle childhood and adolescence, as exhibited by psychosis-spectrum youth, could reflect impairments in optimal specialization. However, these observations are speculative, and the veracity of these patterns will be most accurately captured with longitudinal analyses that encompass a wider age range.

While psychosis has been associated with white matter disruptions, the processes by which these disruptions occur remain unclear. In this study, psychosis-spectrum youth exhibited age-associated abnormalities in RD, the DTI measure associated with myelination,^74, 77, 78^ when compared to typically developing youth in multiple thalamus-prefrontal tracts. Typically developing youth exhibited a u-shaped age-associated trajectory in RD from 14-17 years that was not exhibited by psychosis-spectrum youth, perhaps suggesting that myelination processes are altered in psychosis-spectrum youth during adolescence. In contrast, we did not find any age-associated abnormalities in AD, the DTI measure for axonal density,^74–76^ but we did find age-associated differences in MD, the DTI measure associated with the amount of water content,^79, 80^ when comparing psychosis-spectrum youth to typically developing youth. These findings indicate that abnormal changes in myelination may contribute to disruptions in thalamocortical connectivity in psychosis.

When comparing psychosis-spectrum youth to typically developing youth, we found group differences in GFA in multiple thalamocortical tracts and age-associated abnormalities in GFA in multiple thalamus-prefrontal tracts. Notably, we did not find other significant group differences or age-associated abnormalities with other DSI measures when comparing psychosis-spectrum youth to typically developing youth. Traditionally, DSI measures are thought to be more sensitive and specific to white matter microstructural organization than DTI measures, as DSI allows for multiple directions of diffusion within a voxel.^35^ However, it is possible that higher-resolution imaging is needed to detect group and age-associated differences through DSI measures.^35^ Additionally, because DSI uses model-free metrics (i.e., it does not assume a specific model for diffusion), whereas DTI uses model-based metrics (i.e., it assumes water diffusion occurs along three principal directions), the DSI model may require diffusion measurements in additional directions to those we collected in this study.^96^ Given that DSI measures are thought to be more sensitive and specific to white matter microstructure than DTI measures,^36–38^ the significant age-associated abnormalities that we found with DTI measures could also be due to differences in fibers crossing in psychosis-spectrum youth, rather than alterations in myelination and/or the amount of water in white matter. Future studies using higher resolution imaging may be able to address this possibility.^41^

There are limitations to the present study. We analyzed a cross-sectional data set; therefore, the age-associated abnormalities identified in this study do not reflect within individual change. In the future, longitudinal studies of psychosis-spectrum youth may better identify how the shape and rate of maturation of subject-specific developmental trajectories in psychosis-spectrum youth differ in comparison to typically developing youth.^29, 97, 98^ Moreover, because psychotic symptoms are often dynamic and change over time,^99, 100^ these changes need to be considered when characterizing neurodevelopmental change. Time-varying analytic approaches^101, 102^ could be applied to longitudinal neuroimaging and psychotic symptom data to link psychotic symptoms to specific DWI measures at precise periods of development. Furthermore, although DWI can detect differences in structural connectivity, microstructural differences that result from neurobiological differences in myelination, axonal density, or water content may be better detected with higher-resolution imaging and/or comparisons with myelin-based imaging^103^ or free-water imaging.^104^ Finally, in our analyses, we only examined the whole thalamus as a ROI. Future investigations may look at thalamocortical development in terms of individual thalamic nuclei, to understand whether a particular sub-nucleus (e.g., mediodorsal nucleus), is driving the group- and age-differences seen in our study.

Taken together, our results provide compelling novel evidence for age-associated disruptions of frontal white matter thalamocortical connectivity in psychosis-spectrum youth. In the future, we plan to test the extent to which these findings are observed in longitudinal data sets of youth over the course of development (e.g., the Adolescent Brain and Cognitive Development Study) to better understand how within-subject neurodevelopment contributes to the neurobiological mechanism that underlie psychosis onset.

## Data Availability

All data produced in the present study are available upon reasonable request to the authors

## Notes

### Competing Interest Statement

The authors have declared no competing interest.

### Funding Statement

This study was funded by the NIMH R01 MH129636

### Author Declarations

the Philadelphia Neurodevelopmental Cohort

