## Supplemental Materials for "Age-associated alterations in thalamocortical structural connectivity in psychosis-spectrum youths"

**Supplementary Materials for “Age-associated Alterations in Thalamocortical Structural Connectivity in Psychosis-spectrum Youths”**

***Project Timeline***

This project initially began as Lydia Lewis’ thesis for her undergraduate Neuroscience degree. Dr. Maria Jalbrzikowski served as thesis advisor. The thesis project, *Age-associated alterations in thalamocortical structural connectivity in first episode psychosis,* was registered on the Open Science Framework (OSF) website, in October of 2018 (<https://osf.io/dy36r>). The thesis project was completed in December 2018 and initial results were intriguing but did not survive statistical corrections. To ensure adequate power in our analyses, we decided to work with a larger data set, the publicly available Philadelphia Neurodevelopmental Cohort. We pre-registered the expanded follow-up project in January of 2019 (<https://osf.io/es3cf>) and began analysis in the new cohort. In addition, we slightly altered our hypotheses due to the results obtained from the original project.

***Youth with other psychopathologies and psychosis spectrum youth fail to exhibit age-associated effects in thalamus-prefrontal QA***

For thalamus-lateral prefrontal and orbitofrontal QA, typically developing youth exhibited an inverted u-shaped trajectory, with significant *increases* in QA from 8.2~12 years old, followed by age-associated decreases in QA from 16-23 years. PS and OP failed to exhibit any thalamus-prefrontal QA age-associated effects. For all groups, QA of the thalamus-lateral temporal, thalamus-medial temporal, and thalamus-occipital tracts showed significant age-associated decreases from middle childhood (8.2 years of age) through early adulthood (23 years of age). In the thalamus-parietal tract, TD and OP exhibited a similar linear decrease in QA between 8.2 and 23 years of age, while PS exhibited a linear decrease in this tract between 12.4 and 23 years of age. All groups exhibited age-associated decreases in QA in the white matter tract connecting the thalamus to the supplementary motor cortex beginning in their early teens (~14 years of age) through young adulthood (23 years of age).

***Psychosis spectrum youth fail to exhibit age-associated effects in thalamocortical GFA***

Both TD and OP exhibited age-associated increases in GFA from 8.2~16 years of age in thalamus-prefrontal tracts, while PS failed to show any age-associated effects in these connections. TD also exhibited a similar age-associated increase (8.2-15.3 years) in thalamus-orbitofrontal GFA, while OP and PS youth did not show this pattern. Between 8.2-23 years, TD exhibited a significant age-associated decrease in thalamus-medial temporal GFA; OP and PS both failed to exhibit these effects. OP exhibited an inverted u-shaped trajectory in thalamus-supplementary motor GFA, with a significant age-associated increase in GFA from 8.2-12.4 years, followed by a significant age-associated decrease in GFA from 15.2-23 years. PS and TD failed to exhibit age-associated effects in GFA of the thalamus-supplementary motor tract.

***Psychosis spectrum youth fail to show age-associated effects in thalamus-prefrontal ISO***

In the thalamus-lateral prefrontal, thalamus-medial prefrontal, and thalamus-orbitofrontal tracts, TD youth exhibited significant age-associated decreases in isotropy between ages ~14-23 years. In OP youth, the age range was extended for thalamus-lateral prefrontal and thalamus-orbitofrontal ISO decreases (8.2-23). PS youth failed to exhibit age-associated changes in ISO of thalamus-lateral prefrontal, thalamus-medial prefrontal, or thalamus-orbitofrontal tracts. All groups exhibited significant age-associated linear decreases in ISO in the remaining thalamocortical tracts (thalamus-lateral temporal, thalamus-medial temporal, thalamus-parietal, thalamus-occipital, thalamus-supplementary motor) between ~9 and 23 years of age.

***Both OP and PS failed to exhibit age-associated effects in thalamus-prefrontal RDI***

For thalamus-lateral prefrontal, thalamus-medial prefrontal, and thalamus-orbitofrontal RDI, TD exhibited age-associated decreases from ~14-23 years of age. PS failed to exhibit age-associated effects in RDI of all three observed thalamus-prefrontal tracts, while OP failed to exhibit age-associated affects in RDI of thalamus-lateral prefrontal and thalamus-medial prefrontal tracts. OP showed age-associated decreases in the thalamus-orbitofrontal RDI between 8.2-23 years of age. All groups exhibited linear decreases in RDI for connections between the thalamus and lateral temporal, medial temporal, parietal, occipital, supplementary motor cortices from childhood to early adulthood (~9~23 years).

***PS and OP exhibited age-associated effects in thalamus-prefrontal FA***

TD and OP exhibited increases in FA in three of the observed thalamocortical tracts (thalamus-orbitofrontal, thalamus-parietal, and thalamus-occipital) from 8.2~17 years of age. All groups exhibited significant increases in FA starting at 8.2 years in FA of the thalamus-lateral prefrontal, thalamus-medial prefrontal, and thalamus-supplementary motor tracts. In the thalamus-lateral prefrontal tract, the increase in FA occurred between 8.2~17 years of age in TD and OP, whereas it occurred from 8.2-23 years of age in PS. For all three groups, FA in the thalamus-medial prefrontal tract increased between 8.2~18 years of age. All three groups exhibited significant increases in FA in connections between the thalamus and supplementary motor cortex between 8.2~16 years of age. However, for TD and OP, the increase in FA was followed by a significant decrease in FA in early adulthood (21-23 years of age); this age-associated effect was not observed for PS. TD exhibited a significant increase in FA in the thalamus-medial temporal tract between 10.5-23 years of age; this effect was not observed for OP and PS.

***Psychosis spectrum youth and youth with other psychopathology fail to show normative age-associated decreases in MD in thalamus-frontal tracts***

Typically developing youth and OP exhibited significant decreases in MD from 8.2~16 years of age in the thalamus-parietal tract; however, PS youth failed to exhibit similar age-associated changes. Between 8.2~15 years of age, TD also exhibited age-associated decreases in MD in thalamus-medial prefrontal, -orbitofrontal and -supplementary motor tracts. The decrease in thalamus-supplementary motor MD observed in TD was followed by a significant increase in MD between 17.5-23 years of age. These age-associated effects were absent in OP and PS. Notably, youth with other psychopathology exhibited a u-shaped trajectory in thalamus-occipital MD, with significant *decreases* in MD from 8.2~14.4 years of age, followed by an age-associated increase in MD from 17-23 years of age.

***Psychosis-spectrum youth failed to show age-associated decreases in RD in most thalamocortical tracts***

Typically developing youth exhibited age-associated decreases in RD from middle childhood through adolescence (8.2~16 years of age) in six of the eight thalamocortical tracts (thalamus-lateral prefrontal, thalamus-medial prefrontal, thalamus-orbitofrontal tracts. OP exhibited a similar trend in thalamus-medial prefrontal RD, while PS failed to exhibit any age associated changes in thalamus-frontal RD. TD exhibited a u-shaped trajectory in thalamus-supplementary motor and thalamus parietal RD, with significant decreases from 8.2~15 years of age, and significant increases from ~17-23 years of age. OP exhibited a significant increase in th alamus-parietal RD between 8.2-15.7 years of age, while PS failed to exhibit significant age effects in the thalamus-supplementary motor and thalamus-parietal tracts. OP exhibited a u-shaped trajectory in thalamus-occipital RD, with a significant decrease in RD between 8.2-14.5 years of age and a significant between 17.1-23 years of age. TD exhibited a significant linear increase in thalamus-medial temporal RD from 8.2-23 years of age; OP and PS both failed to exhibit this age effect.

**Psychosis spectrum youth failed to exhibit age-associated changes in AD**

Typically developing youth exhibited age-associated decreases in AD in three of the eight thalamocortical tracts: thalamus-supplementary motor, thalamus-occipital, and thalamus-parietal; OP exhibited an age-associated decrease in AD in only the thalamus-parietal tract, while PS did not exhibit any age-associated changes in AD. For TD, age-associated changes occurred from 8.2~16 years of age in the thalamus-supplementary motor and thalamus-parietal tracts. From 8.2-23 years of age, linear decreases in AD occurred for TD in the thalamus-occipital tract and occurred for OP in the thalamus-parietal tract.

**Supplemental Table 1.** Cortical regions of interest (ROI) used as tractography targets in this investigation. Next to each cortical ROI is a list of the regions in the Freesurfer Desikan-Killiany atlas that were merged to create each ROI. The insula was not included in our study because initial tractography did not identify tracts between the thalamus and insula. A three-dimensional visualization of these ROIs is presented in Figure 1.

| **ROI** | **FreeSurferDKT regions included:** |
| --- | --- |
| Orbitofrontal Cortex (OFC) | Pars orbitalis, medial orbitofrontal cortex, lateral orbitofrontal cortex |
| Medial Prefrontal Cortex (MPFC) | Caudal anterior cingulate, rostral anterior cingulate, superior frontal gyrus |
| Lateral Prefrontal Cortex (LPFC) | Parts triangularis, rostral middle frontal gyrus, pars opercularis, caudal middle frontal gyrus |
| Sensorimotor Cortex (SMC) | Precentral gyrus, postcentral gyrus, paracentral lobule |
| Parietal Cortex (PC) | Inferior parietal cortex, supramarginal gyrus, precuneus cortex, posterior cingulate cortex, isthmus cingulate, superior parietal cortex |
| Medial Temporal Cortex (MTC) | Entorhinal cortex, parahippocampal gyrus, fusiform gyrus |
| Lateral Temporal Cortex (LTC) | Transverse temporal cortex, superior temporal gyrus, inferior temporal gyrus, middle temporal gyrus |
| Occipital Cortex (OCC) | Pericalcarine cortex, lingual gyrus, lateral occipital cortex, cuneus cortex |
